# Over one year of survival difference between distinct cohorts of glioblastoma patients

**DOI:** 10.1101/2023.03.20.23287451

**Authors:** Michael TC Poon, Peter YM Woo, Aya El Helali, Paul M Brennan

**Affiliations:** Department of Clinical Neurosciences, Royal Infirmary of Edinburgh, Edinburgh, United Kingdom; Translational Neurosurgery, Centre for Clinical Brain Sciences, University of Edinburgh, United Kingdom; Usher Institute, University of Edinburgh, Edinburgh, United Kingdom United Kingdom; Department of Neurosurgery, Prince of Wales Hospital, Hong Kong; Department of Clinical Oncology, School of Clinical Medicine, University of Hong Kong, Hong Kong

**Author notes:** **Corresponding author** Dr. Michael Tin Chung Poon, Department of Neurosurgery, Department of Clinical Neurosciences, Royal Infirmary of Edinburgh, 50 Little France Crescent, Edinburgh, EH16 4SA, United Kingdom. **Author contributions** Conceptualisation: MTCP; methodology: MTCP, PMB; formal analysis: MTCP; data curation: MTCP, PYMW, AEH, PMB; manuscript draft: MTCP; manuscript review and editing: MTCP, PYMW, AEH, PMB.

## Abstract

Understanding the differences in outcomes based on study and patient characteristics can inform better research designs and improve the interpretation of translational glioblastoma studies. This study compared the clinical features and median overall survival (mOS) of 2,203 patients from two genomic cohorts (The Cancer Genome Atlas [TCGA] and the Chinese Glioma Genome Atlas [CGGA]) and two consecutive regional cohorts (Hong Kong [HK] and Southeast [SE] Scotland) with histologically confirmed glioblastoma. Differences in clinical characteristics reflected the distinct patient selection criteria for surgery. While the mOS were similar, the mOS of those who completed temozolomide chemoradiotherapy were 35.2 months in SE Scotland, 22.2 in HK, and 14.1 in the TCGA cohort. Survival functions were comparable in the multivariable survival analysis. Our findings highlight how a lack of clinical data impedes the translational value of research. Therefore, there is a need to develop common data elements to derive meaningful conclusions.

## Introduction

Glioblastoma is an incurable brain tumour; only 4% of people survive more than 5 years after diagnosis.^1^ Standard-care treatment has remained unchanged for two decades.^2^ With an increasing understanding of tumour biology, several molecular markers have been implicated as critical for predicting patient survival.^3^ These biomarkers can support precision medicine and stratification for therapeutic interventions. Establishing the utility of biomarkers requires a thorough understanding of the study and target patient population. The generation of research data relevant to patients is the foundation of translational and clinical studies. A robust study design underpins the quality of such data and can affect the interpretation of findings. ‘Omics’ studies that rely on tumour samples require informed patient consent, inevitably introducing selection bias. In addition, ethnic composition and healthcare settings can introduce outcome variations between cohorts. The validity of an observation requires reproducibility demonstrable in distinct geographical settings. While investigators are aware of these considerations, the extent of their impact on outcome studies has not been described, not least because of the logistical and technical challenges of curating the necessary datasets.

Understanding the degree of variability and inter-study survival outcomes based on study and patient characteristics can inform better translational glioblastoma research methodologies. Therefore, we combined two consecutive regional cohorts and two genomic cohorts of newly diagnosed glioblastoma patients to describe survival outcomes stratigied by study design and treatment strategies.

## Methods

### Study setting and patients

This retrospective cohort study included consecutive patients aged ≥ 18 years who underwent surgery for newly diagnosed glioblastoma in Southeast (SE) Scotland^4^ [Apr 2012-May 2020] and Hong Kong (HK)^5^ [Jan 2006-Dec 2019]. In addition, we also studied newly diagnosed glioblastoma patients aged ≥ 18 years from The Cancer Genome Atlas^6^ (TCGA) and Chinese Glioma Genome Atlas^7^ (CGGA). Full descriptions of the cohorts are available in the index publications. Data collection in the regional cohorts was approved by the Southeast Scotland Research Ethics Committee (reference: 17/SS/0019) and Hong Kong Hospital Authority Institutional Review Board (reference: KC/KE-18-0262/ER-4). Study entry was the date of primary surgery that yielded a histological diagnosis of glioblastoma. Specialist neuro-oncology services subsequently provided follow-up care for patients within consecutive cohorts. Sources of information included electronic health records, histopathology reports, and radiological images. The censoring dates for survival data were the 16th of January 2021 and the 31st of October 2021 for SE Scotland and HK, respectively.

### Variables

The different study designs required data harmonisation. We categorised the preoperative Karnofsky Performance Status (KPS) score as ≥ 70 or <70. The extent of resection included biopsy, subtotal resection, and gross total resection. The SE Scotland cohort used postoperative contrast-enhanced MRI within 72 hours, and the HK cohort used the surgeons’ intraoperative assessment to determine the extent of resection. Standard chemoradiotherapy denotes radiotherapy plus concomitant and adjuvant temozolomide.^2^ Tumour O^6^-methylguanine-DNA-methyltransferase (*MGMT*) gene promoter methylation status was determined using pyrosequencing and methylation-specific polymerase chain reaction in SE Scotland and HK, respectively. Overall survival (OS) was defined as the period from the date of the primary operation to the date of death.

### Bias

Chronological bias may affect our study because of the long study period. Changing diagnostic practices over the study period may have systematically affected patient characteristics. For example, increased use of radiological imaging may identify more patients with no or mild symptoms and better performance status. In addition, treatment options have changed; HK and SE Scotland incorporated temozolomide into their hospital formularies in 2010 and 2007, respectively. To reduce these biases in our multivariable analyses, we analyzed patients diagnosed after 2010.

### Statistical analyses

We used descriptive statistics to determine the patient characteristics in the included cohorts. Kaplan-Meier curves were used to visualise survival functions in different settings. Graphs were stratified by study setting (consecutive vs. genomic), data source (SE Scotland, HK, TCGA, and CGGA), *MGMT* methylation status, and patients who completed standard chemoradiotherapy. To include more comparable cohorts, we presented the survival curves of patients with *IDH-1* wildtype glioblastoma diagnosed after the 31st of Dec 2009 who completed standard chemoradiotherapy after surgery. We determined the median overall survival (mOS) using the Kaplan-Meier method. We refrained from using comparative statistics because the cohorts were not directly comparable. To investigate any difference in OS between SE Scotland and HK, we used complete-case multivariable Cox regression adjusted for age, preoperative KPS, extent of resection, *MGMT* status, and oncological therapy.

## Results

### Patient cohorts

A total of 2,203 patients with newly diagnosed glioblastoma were identified, including 414 from SE Scotland, 1,033 from HK, 510 from TCGA, and 246 from CGGA. Follow-up data were not available for 18 patients (Table 1). The total follow-up time was 3,011 person-years with a median follow-up of 0.93 (interquartile range: 0.47-1.65) years. The demographic and clinical characteristics of patients are presented in Table 1. When information was available, 75.1% of the patients had preoperative KPS ≥ 70, and 25.8% had gross total tumour resection. Among the patients, 12.9% did not receive postoperative oncological treatment. CGGA had a younger cohort with more *IDH-1* mutant and *MGMT* methylated tumours (Table 1). Across the cohorts, 561 (25.5%) patients (62 in SE Scotland, 289 in HK, 210 in TCGA) completed standard adjuvant temozolomide chemoradiotherapy (Supplementary Table 1).

**Table 1.**
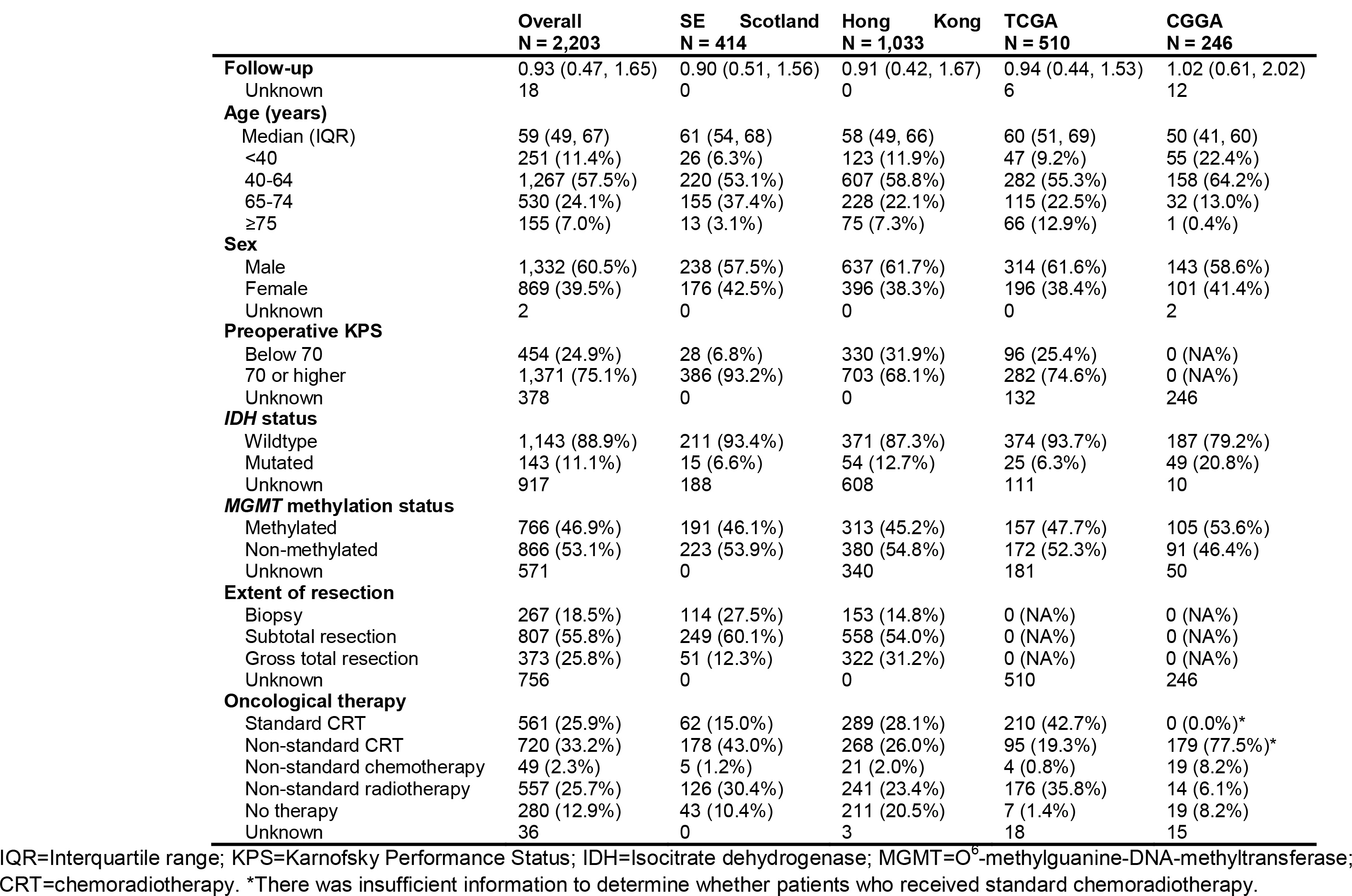
Clinical and treatment characteristics of 2,203 patients from four cohorts.

### Overall survival by cohort setting and tumour characteristics

The mOS of the genomic and consecutive cohorts were 13.1 (95% confidence interval [CI] 12.2-14.0) months and 11.4 (95%CI 10.8-12.1) months, respectively (Figure 1A). In individual cohorts, mOS was 13.6 months (95%CI 12.4-14.5) in TCGA, 12.4 (95%CI 10.9-14.4) in CGGA, 11.6 (95%CI 10.8-12.6) in HK, and 10.9 (95%CI 10.1-12.3) in SE Scotland (Figure 1B). Among those diagnosed with *IDH-1* wildtype glioblastoma after the 31st of December 2009 and who received standard chemoradiotherapy, mOS was 35.2 months (95%CI 27.1-50.2) in SE Scotland, 22.2 (95%CI 19.0-25.5) in HK, and 14.1 months (95%CI 11.8-17.9) in TCGA (Figure 1C). When these patients were stratified by *MGMT* status, those with *MGMT* methylated tumours had longer mOS (36.2 months in SE Scotland, 26.8 months in HK, and 16.8 months in TCGA) compared to those with *MGMT* unmethylated tumours (24.5 months in SE Scotland, 17.0 months in HK, and 12.6 months in TCGA) (Figure 1D).

**Figure 1.**
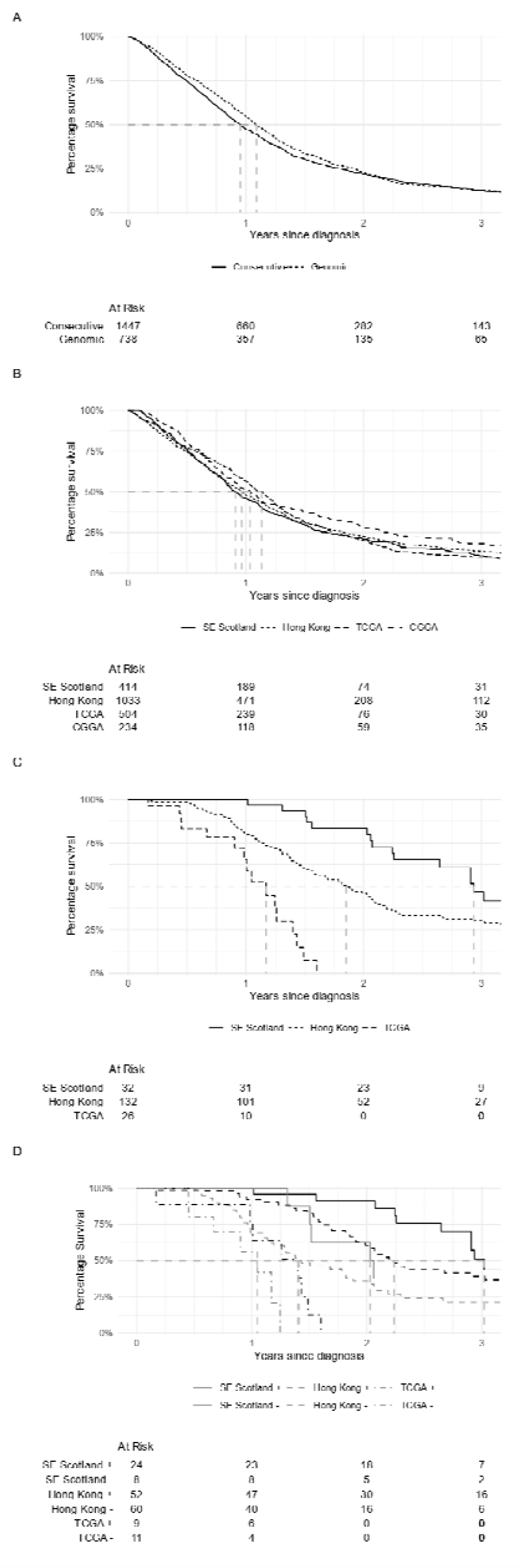
Survival functions of patients diagnosed with glioblastoma in different contexts (A) Survival functions of consecutive cohorts (SE Scotland and Hong Kong) and genomic cohorts (TCGA and CGGA). (B) Individual survival functions of included cohorts. (C) Survival function by cohort of patients diagnosed with *IDH*-wildtype glioblastoma after the 31^st^ Dec 2009 who completed standard chemoradiotherapy. (D) Same patients as (C) with stratification by *MGMT* promoter methylation status where ‘+’ denotes methylated and ‘-’ denotes unmethylated tumours.

### Association between study cohort and overall survival

A total of 1,023 patients from SE Scotland (N=414) and HK (N=609) with 1,446.6 person-years of follow-up and 901 deaths were included in the survival analysis. Multivariable survival analysis across all four cohorts was not feasible because of the missing key variables. The multivariable Cox regression model did not demonstrate survival differences among patients treated in HK compared to those treated in SE Scotland (hazard ratio 0.93; 95%CI 0.80-1.08; p=0.30) (Table 2).

**Table 2.** Multivariable Cox regression of 1,023 patients diagnosed on or after the 1^st^ of Jan 2010 in Southeast (SE) Scotland and Hong Kong

| Variables | HR | 95% CI | p-value |
| --- | --- | --- | --- |
| *Cohort* |  |  |  |
| SE Scotland | ref | - | - |
| Hong Kong | 0.93 | 0.80-1.08 | 0.3 |
| *Age* |  |  |  |
| Each year | 1.01 | 1.01-1.02 | <0.001 |
| *Preoperative KPS* |  |  |  |
| Below 70 | ref | - | - |
| 70 or higher | 0.87 | 0.73-1.03 | 0.11 |
| *Extent of resection* |  |  |  |
| Biopsy | ref | - | - |
| Subtotal resection | 0.67 | 0.56-0.79 | <0.001 |
| Gross total resection | 0.63 | 0.51-0.77 | <0.001 |
| *MGMT promotor status* |  |  |  |
| Non-methylated | ref | - | - |
| Methylated | 0.64 | 0.56-0.74 | <0.001 |
| *Oncological therapy* |  |  |  |
| No therapy | ref | - | - |
| Non-standard chemotherapy | 0.26 | 0.15-0.45 | <0.001 |
| Non-standard radiotherapy | 0.36 | 0.28-0.45 | <0.001 |
| Non-standard CRT | 0.21 | 0.17-0.26 | <0.001 |
| Standard CRT | 0.09 | 0.07-0.12 | <0.001 |

KPS=Karnofsky Performance Status; MGMT= O<sup>6</sup>-methylguanine-DNA-methyltransferase; CRT=chemoradiotherapy

## Discussion

Comparing 2,203 patients from four geographically distinct glioblastoma cohorts with different study designs, we found inconsistent data availability of common clinical characteristics. In particular, there were differences in data availability regarding functional status, extent of resection, and oncological treatment between the cohorts. In addition, the genomic cohorts showed better unadjusted survival outcomes than consecutive regional cohorts. However, survival in patients who completed chemoradiotherapy was higher in the consecutive cohorts than in the genomic cohort. There was no difference in the hazard of death between the HK and SE Scotland cohorts when accounting for clinical variables.

We demonstrated substantial differences in survival among patients who completed standard chemoradiotherapy between our cohorts. This finding has implications for the interpretation of translational research using various patient selection methods. Demonstrating the relevance of biological pathways in human diseases is essential for translational research. Preclinical studies often validate their findings using survival analysis from human genomic resources.^8^ Our findings confirmed strong associations between clinical variables and survival that can confound univariable survival analysis, resulting in misleading results. Harmonising cohort data on vital clinical characteristics can be difficult because of different study designs and diverse clinical practices. These challenges in data availability call for the establishment of common data elements to facilitate comparable clinical glioblastoma research. National or regional cancer registries can use this set of common data elements to evaluate the diagnostic and treatment patterns. Both clinical research and cancer registries should classify brain tumours to reflect the understanding of molecular profile defined by the World Health Organisation’s (WHO) latest classification.^9^

### Strengths & limitations

We combined four cohorts to demonstrate survival functions based on different study settings and patient selection. Our consecutive patient cohorts allowed us to present the clinical characteristics and outcomes with minimal selection bias. The limitations of our study include the lack of standardised variable definitions in the cohorts. It is essential for future comparative studies to specify the data ascertainment method. We could not include all cohorts in our multivariable analysis because of data availability. Such an analysis would provide further information regarding the biases associated with the study setting. We were unable to review patients with glioblastoma according to the latest 2021 WHO definition that required *IDH*-*1* wildtype determination. However, such molecular data was likely missed at random, so our findings were unlikely to be affected by this.^9^

## Conclusions

Translational glioblastoma research using patient-derived tumour tissues must incorporate clinical variables when evaluating the outcomes. Common data element documentation allows for adjusted analyses and investigation of differences in outcomes between patient cohorts. Ensuring an adequate description of the study cohort is vital for translational, biomarker, and prognostic studies.

## Data Availability

Data from the consecutive cohorts that support the findings of this study are available on request from the corresponding author MTCP. The data are not publicly available due to clinical data governance agreement. Data from TCGA (https://portal.gdc.cancer.gov/) and CGGA (http://www.cgga.org.cn/download.jsp) are available in their respective repositories.

---

**Supplementary Table 1.** Clinical characteristics of 561 patients who completed standard chemoradiotherapy after surgery

| **Characteristic** | **Overall**<br>**N = 561** | **SE Scotland**<br>**N = 62** | **Hong Kong**<br>**N = 289** | **TCGA**<br>**N = 210** |
| --- | --- | --- | --- | --- |
| **Age (years)** |  |  |  |  |
| <40 | 84 (15.0%) | 12 (19.4%) | 47 (16.3%) | 25 (11.9%) |
| 40-64 | 368 (65.6%) | 42 (67.7%) | 194 (67.1%) | 132 (62.9%) |
| 65-74 | 86 (15.3%) | 8 (12.9%) | 46 (15.9%) | 32 (15.2%) |
| ≥75 | 23 (4.1%) | 0 (0.0%) | 2 (0.7%) | 21 (10.0%) |
| **Sex** |  |  |  |  |
| Male | 340 (60.6%) | 32 (51.6%) | 178 (61.6%) | 130 (61.9%) |
| Female | 221 (39.4%) | 30 (48.4%) | 111 (38.4%) | 80 (38.1%) |
| **Preoperative KPS** |  |  |  |  |
| Below 70 | 101 (19.2%) | 2 (3.2%) | 64 (22.1%) | 35 (20.1%) |
| 70 or higher | 424 (80.8%) | 60 (96.8%) | 225 (77.9%) | 139 (79.9%) |
| Unknown | 36 | 0 | 0 | 36 |
| **IDH mutation** |  |  |  |  |
| Wildtype | 323 (86.6%) | 32 (82.1%) | 133 (82.1%) | 158 (91.9%) |
| Mutated | 50 (13.4%) | 7 (17.9%) | 29 (17.9%) | 14 (8.1%) |
| Unknown | 188 | 23 | 127 | 38 |
| **MGMT methylation status** |  |  |  |  |
| Methylated | 265 (57.6%) | 48 (77.4%) | 136 (57.4%) | 81 (50.3%) |
| Non-methylated | 195 (42.4%) | 14 (22.6%) | 101 (42.6%) | 80 (49.7%) |
| Unknown | 101 | 0 | 52 | 49 |
| **Extent of resection** |  |  |  |  |
| Biopsy | 40 (11.4%) | 6 (9.7%) | 34 (11.8%) | 0 (NA%) |
| Subtotal resection | 197 (56.1%) | 45 (72.6%) | 152 (52.6%) | 0 (NA%) |
| Gross total resection | 114 (32.5%) | 11 (17.7%) | 103 (35.6%) | 0 (NA%) |
| Unknown | 210 | 0 | 0 | 210 |

KPS=Karnofsky Performance Status; IDH=Isocitrate dehydrogenase; MGMT= O<sup>6</sup>-methylguanine-DNA-methyltransferase. There was insufficient information to determine whether patients who received chemotherapy and radiotherapy received standard chemoradiotherapy in the CGGA cohort.

